# Partitioning and probe-based quantitative PCR assays for the wastewater monitoring of *Mycobacterium tuberculosis* Complex, *M. tuberculosis*, and *M. Bovis*

**DOI:** 10.1101/2024.12.18.24319238

**Authors:** Tram B. Nguyen, Elisabeth Mercier, Chandler H. Wong, Nada Hegazy, Md Pervez Kabir, Emma Tomalty, Felix Addo, Leonor Ward, Elizabeth Renouf, Shen Wan, Yassen Tcholakov, Stéphanie Guilherme, Robert Delatolla

## Abstract

Three new probe-based quantitative PCR assays were designed based on Chae *et al*. (2017), *Pérez-Osorio et al.* (2012), and Sales *et al*. (2012) to quantitate *Mycobacterium tuberculosis* complex (MTBC) species, *M. tuberculosis* (MTB), and *M. bovis* (MB) in wastewater targeting genomic regions *rv0577,* RD9, and the deletion of RD4, respectively. The assays were validated for specificity using four Mycobacterial species, including two MTBC species and two non-tuberculosis Mycobacteria species, and endogenous wastewater samples from Ottawa, Ontario, Canada, Mumbai, India, and a remote Northern Indigenous community in Nunangat with known ongoing tuberculosis cases or outbreaks. The three assays demonstrate high sensitivity and are suitable for use in wastewater. Partitioning experiments performed on endogenous MTBC and MTB in collected wastewaters from Mumbai, India with known tuberculosis outbreaks show that the targeted genomic regions of *rv0577* (MTBC) and RD9 (MTB) used to quantitate human tuberculosis infection predominately partition to solids fraction of wastewaters. The partitioning results of this study, in combination with the presented probe-based PCR assays, provide guidance on how to best enrich wastewaters and rapidly and economically quantify tuberculosis with high specificity and sensitivity in wastewaters.

**GRAPHICAL ABSTRACT:** 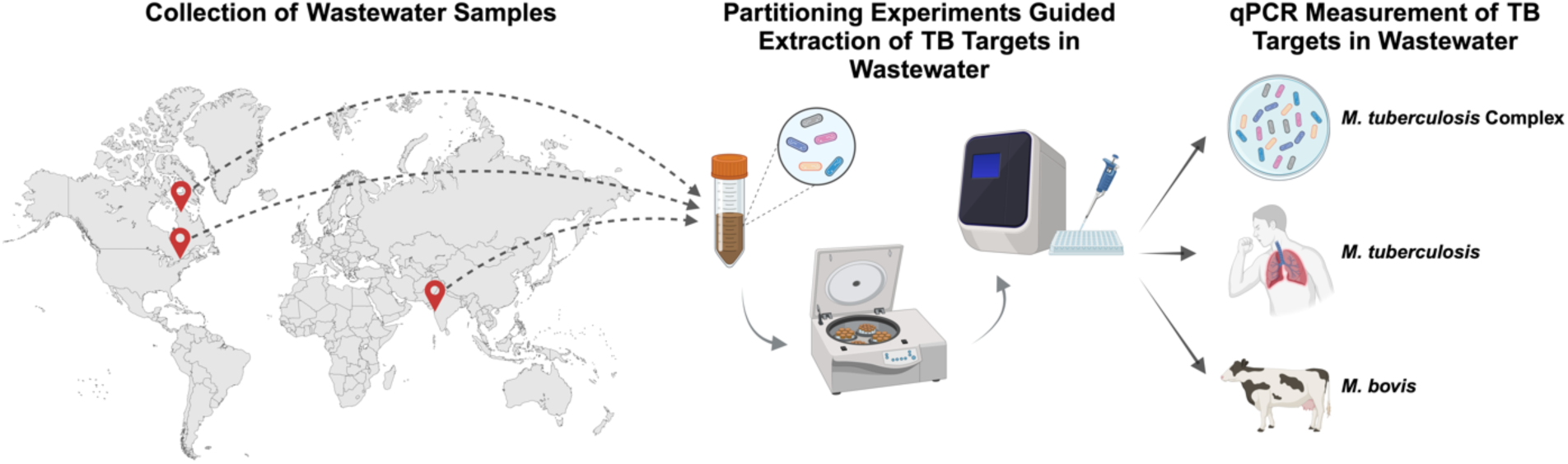

## INTRODUCTION

Tuberculosis (TB) infections were ranked the most lethal infectious disease globally in 2023, resulting in over 1.25 million fatalities. Approximately 10.8 million people became ill with TB worldwide in 2023, with most cases occurring in Southeast Asia (45%) and Africa (24%). The 30 top TB-burdened countries account for 87% of all incident cases globally, including India (26%), Indonesia (10%), and China (6.8%) (World Health Organization, 2024). TB is caused by *Mycobacterium tuberculosis* complex (MTBC) species, a group of closely related mycobacterial strains capable of causing infections in various animal species, including humans. MTBC species share highly conserved genomes that result in potential zoonotic TB (zTB) infection potential across animal species (Bayraktar *et al*., 2011; Lawn and Zumla, 2011; Langer and LoBue, 2014; Jia *et al*., 2017; Romha *et al*., 2018; Correa-Macedo, Cambri and Schurr, 2019; Kanabalan *et al*., 2021; Kock *et al*., 2021). Individuals who are frequently in close contact with infected animals, such as livestock, are hence at risk of zTB infection of other MTBC species, such as *M. bovis* (MB), *M. orygis,* and *M. caprae* (Olea-Popelka *et al*., 2017; Duffy *et al*., 2024; Neil M. Ampel, 2024). Although zTB infection is possible in humans, *M. tuberculosis* (MTB) is the primary MTBC species responsible for human infection (Gagneux, 2012)[14]. Human TB infection most often begins when an individual inhales droplets or bodily fluids containing viable MTBC bacteria from an infected human host (Sakamoto, 2012), with 90-95% of exposure events resulting in latent TB infection (LTBI) (Greenaway *et al*., 2011). LTBI is a state of persistent immune response to MTB antigens, with no evidence of clinically manifest active TB (World Health Organization, 2019). It is estimated that up to one-third of the world population is latently infected with MTB (World Health Organization, 2019).

In 2022, Canada documented 1,971 incident active TB cases, with a rate of active TB infections being 5.1 cases per 100 000 individuals. Within this group, 74.5% of documented cases had occurred in foreign-born individuals, at 12.3 cases per 100 000 individuals. The lowest rates of active TB occurrence in Canada are in non-Indigenous, Canadian-born individuals, at a rate of 0.3 cases per 100 000 individuals (Public Health Agency of Canada, 2024). Indigenous populations in Canada have the highest rate of incident active TB cases, at a rate of 16.6 cases per 100 000 individuals. (Statistics Canada, 2019). The Inuit population of Nunangat (or ᐃᓄᐃᑦ ᓄᓇᖓᑦ in Inuktitut) is located in Canada’s Arctic region and is home to 69.1% of the Inuit population in Canada (Statistics Canada, 2022). The Nunangat population is disproportionately affected by TB, with the average incidence rate being more than 290 times higher than non-Indigenous, Canadian-born individuals (Patterson, Flinn and Barker, 2018). The Nunangat population experiences systemic inequality and inequity issues such as crowded housing, food insecurity, lack of access to adequate healthcare, water insecurity, and unemployment as compared to non-Indigenous Canadians. These social determinants of health are known to exasperate TB infection rates (Clark, Riben and Nowgesic, 2002; Tuite *et al*., 2017; Patterson, Flinn and Barker, 2018; Sinha *et al*., 2019; Cassivi *et al*., 2023). Despite an overall low prevalence of TB in Canada, these specific communities and populations are still disproportionately impacted by TB.

Currently, the most common method for identifying active TB cases in Nunangat is community-wide screening, which is difficult and expensive to conduct due to the lack of health infrastructure available. Since the communities of this region are often remote, issues such as limited available space and housing pose significant barriers to obtaining the necessary health infrastructures to conduct effective screening (Patterson, Flinn and Barker, 2018). Furthermore, TB infection identification in Nunangat traditionally first requires an infected person to present with symptoms and then volunteer to complete clinical diagnosis, followed by reporting to public health officials. Indigenous communities, including Inuit populations, in Nunangat and across Canada have experienced a well-documented, complex, and traumatic medical history related to TB testing. This history has resulted in medical testing hesitancy and, hence, often an under-representation of individuals with TB in Indigenous communities by public health agencies across the country (Vachon, Gallant and Siu, 2018; Basta and de Sousa Viana, 2019; Hick, 2019; Jetty, 2020).

Wastewater and environmental monitoring (WEM) provides an anonymous and non-invasive means to potentially provide Nunangat communities with an early warning system of TB illness and transmission (Williams *et al*., 2024). MTB has been detected in stool from infected patients using both culture- and molecular-based methods (Jensen, 1954; Greenberg and Kupka, 1957; Oramasionwu *et al*., 2013; Kokuto *et al*., 2015; Mesman *et al*., 2019; Kesarwani *et al*., 2022; Mtetwa *et al*., 2022a). Since the early 20^th^ century, MTB has been detected in wastewater, from the detection in sanatorium wastewater to, more recently, wastewater monitoring in water recovery and reuse facilities (WRRF) in high-prevalence regions (Greenberg and Kupka, 1957; Mtetwa *et al*., 2022a). Wastewater from facilities housing TB patients, such as sanatoria, has been used to detect viable bacteria and successfully used as a vector to infect animals such as Guinea pigs with TB (Brown, Petroff and Heise, 1916; Kroger and Trettin G, 1951; Jensen, 1954; Greenberg and Kupka, 1957). After the onset of the COVID-19 pandemic, there has been a pronounced resurgence in WEM to track disease incidence and provide early identification of outbreaks and disease transmission in communities around the world (Schmidt, 2020; Robins *et al*., 2022). In particular, WEM has been expanded to monitor not only SARS-CoV-2, but also influenza A & B, respiratory syncytial virus (RSV), mpox (formerly known as monkeypox), human metapneumovirus (HMPV), parainfluenza (PIV), norovirus GII, rotavirus, adenovirus group F, enterovirus D68, *Candida auris*, poliovirus, hepatitis A virus (HAV) and others (Mercier *et al*., 2022; Tedcastle *et al*., 2022; Boehm, Hughes, *et al*., 2023; Boehm, Wolfe, *et al*., 2023; Kitakawa, Kitamura and Yoshida, 2023; Sherchan *et al*., 2023).

Similar to clinical diagnoses, many current protocols used to detect MTB in wastewater have utilized culture-based methods. Newer methodologies, such as polymerase chain reaction (PCR) and shotgun sequencing, have since been used (Mtetwa *et al*., 2022b). In clinical settings, the IS6110 PCR target is utilized for MTB detection via PCR. However, this PCR target lacks specificity for wastewater applications due to non-specific hybridization to extraneous microbial species found in wastewater (Coros, DeConno and Derbyshire, 2008; Müller, Roberts and Brown, 2015). More recently, successful surveillance of MTB in wastewater has been demonstrated using conventional non-probe-based PCR and intercalating dye-based digital droplet PCR (ddPCR) in regions of Africa with high TB incidence (Mtetwa *et al*., 2022a). Intercalating dye-based PCR assays are known to demonstrate lower specificity and sensitivity and are more susceptible to false positive results and non-specific binding to other organisms compared to probe-based quantitative (qPCR) assays (Navarro *et al*., 2015). In order to meet its potential as an early warning system for TB infection in Northern Indigenous communities, WEM TB methods require highly specific assays due to the biologically diverse nature of wastewater and high genomic homology across mycobacterial species, highly sensitive assays to ensure detection at the onset of infection to ensure detection of all TB-infection cases in the community (McEvoy *et al*., 2012). Currently, there is a lack of available probe-based qPCR assays that demonstrate the sensitivities and specificities required for wastewater applications [53]. As a result, we developed and validated wastewater probe-based qPCR assays to detect and quantify not only MTB but also MTBC and MB in both low- and high-incidence municipal wastewater from Ottawa, Ontario, Canada, Mumbai, India, and a Nunangat, Northern Indigenous community with known ongoing TB cases or outbreaks. The developed assays are specific to their respective targets and were validated to rule out non-specific hybridization to extraneous microbial species. The three probe-based qPCR assays were developed based on existing PCR assays (Pérez-Osorio *et al*., 2012; Araújo *et al*., 2014; Sales *et al*., 2014; Chae *et al*., 2017). An assay targeting the *rv0577* region is used to detect and quantitate MTBC species, the region of difference 9 (RD9) is targeted to detect and quantitate MTB, and the RD4 deletion is targeted to detect and quantitate MB. The MTBC *rv0577* and MTB RD9 assays in this study were also used to determine the partitioning behaviour of endogenous MTBC and MTB related to human TB disease in municipal influent wastewater and primary clarified sludge to guide best practices related to the enrichment and concentration of these genetic targets and the application for WEM for TB monitoring. This research provides the necessary information to implement sensitive and specific TB WEM, and hence also provides a pathway towards non-invasive and economical TB monitoring in Northern Indigenous communities in Canada.

## MATERIALS AND METHODS

### qPCR Assay Design

#### Assay design

The MTBC *rv0577* qPCR assay is designed to measure all MTBC species, including MTB and MB, and is a modified version of the Chae *et al*. (2017) *rv0577* assay (Chae *et al*., 2017). The MTB RD9 assay is designed to specifically quantitate MTB and is a modified version of the Pérez-Osorio *et al*. (2012) RD9 assay (Pérez-Osorio *et al*., 2012). The MB RD4-deletion assay is designed to quantitate MB and is a modified version of the Sales *et al*. (2017) Mb.400 assay (Table 1) (Sales *et al*., 2014; Ru *et al*., 2017). The virulence-associated gene, *rv0577,* is found explicitly within the MTBC and used to confirm MTB and MB detections (Gu *et al*., 2016; Chae *et al*., 2017; Qasim, Hameed and Shehzad, 2023). The RD9 intergenic region is targeted within the MTB RD9 assay and is found between the *rv2072c* and *rv2073c* genes that is specific to MTB and not found in other MTBC species (except *M. canetti*) (Chae *et al*., 2017). Other TB-causing mycobacteria and non-TB mycobacteria (NTM) lack the RD9 intergenic region, allowing for molecular differentiation of MTB from other species of MTBC, such as *M. africanum*, *M. orygis, and* MB (Brosch *et al*., 2002; Huard *et al*., 2003; Smith *et al*., 2006; Teo *et al*., 2013). MB lacks the RD4 intergenic region compared to other closely related MBTC species, such as MTB, allowing for molecular differentiation from other species of MTBC (Ru *et al*., 2017; Kapalamula *et al*., 2021). As a result, the MB RD4-deletion assay is used to measure the deletion of the RD4 region in MB. The existing referred PCR products were modified using Primer3Plus (https://www.primer3plus.com) to produce probe-based qPCR assays and subsequently aligned using MUSCLE with sequences of commonly referred MTB variants (H37Rv and HN878), MTBC species (*M. canettii*, *M. africanum*, and *M. microti*), commonly referred MB variants (Mb3602, Danish 1331, bacille Calmette-Guérin (BCG)), and commonly referred NTM species (*M. avium hominissuis, M. kansasii, and M. intracellulare*) to determine *in silico* assay specificity (Figure 1) using genome sequences obtained from GenBank (National Institutes of Health) (Edgar, 2004). The assays employed DNA primers and probes with a 5’-FAM reporter molecule, a 3’ minor groove binder, and a nonfluorescent quencher (Applied Biosystems Integrated).

**Figure 1.**
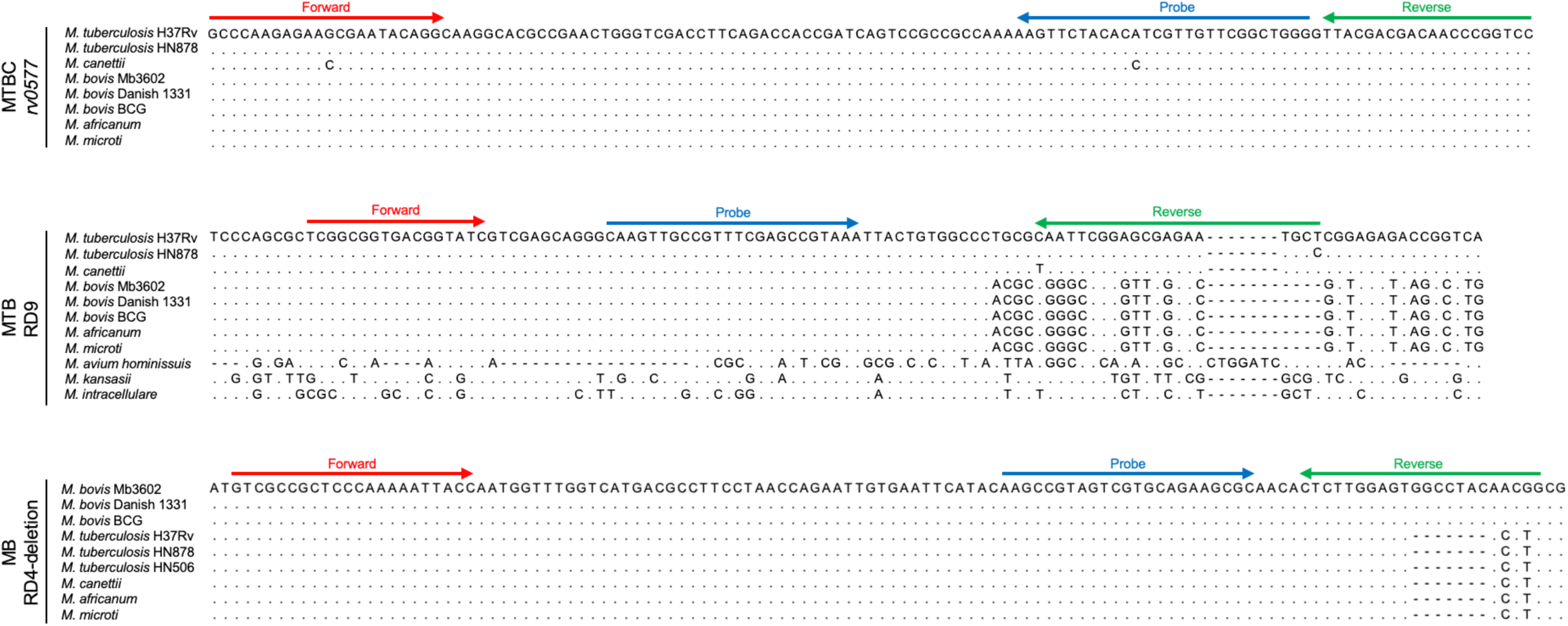
Alignment of primers and probes with mycobacterial DNA genome sequences. The primers and probe for the MTBC rv0577, MTB RD9, and MB RD4-deletion real-time qPCR assays are aligned with the targeted DNA sequence within several mycobacterial species. Mycobacterial strains: *M. tuberculosis* H37Rv (NC_000962), *M. tuberculosis* HN878 (AP018036), *M. canettii* (HE572590), *M. bovis* (NZ_CP096843), *M. bovis* (CP039850), *M. bovis* BCG (AM408590), *M. africanum* (FR878060), MTB *microti* (NZ_LR882496), *M. avium hominissuis* (CP018019), *M. kansasii* strain (CP006835), *M. intracellulare* (CP085945). Dots represent conserved nucleotides, and dashes represent nucleotide gaps.

**Figure 2.**
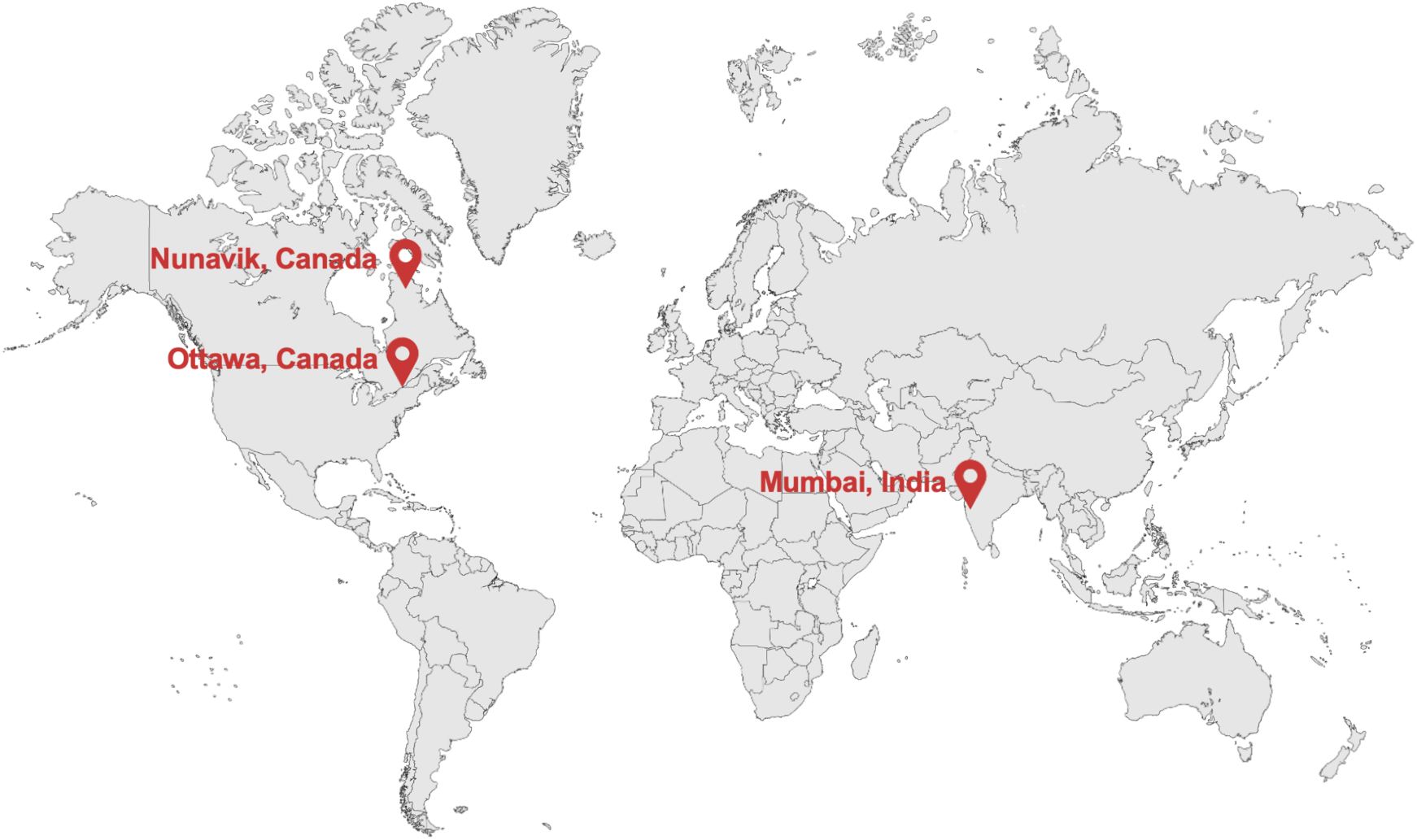
Sampling locations. Red location markers locate Ottawa, Mumbai, and the Nunangat, Northern Indigenous Community, where wastewater samples containing endogenous TB targets were collected.

**Figure 3.**
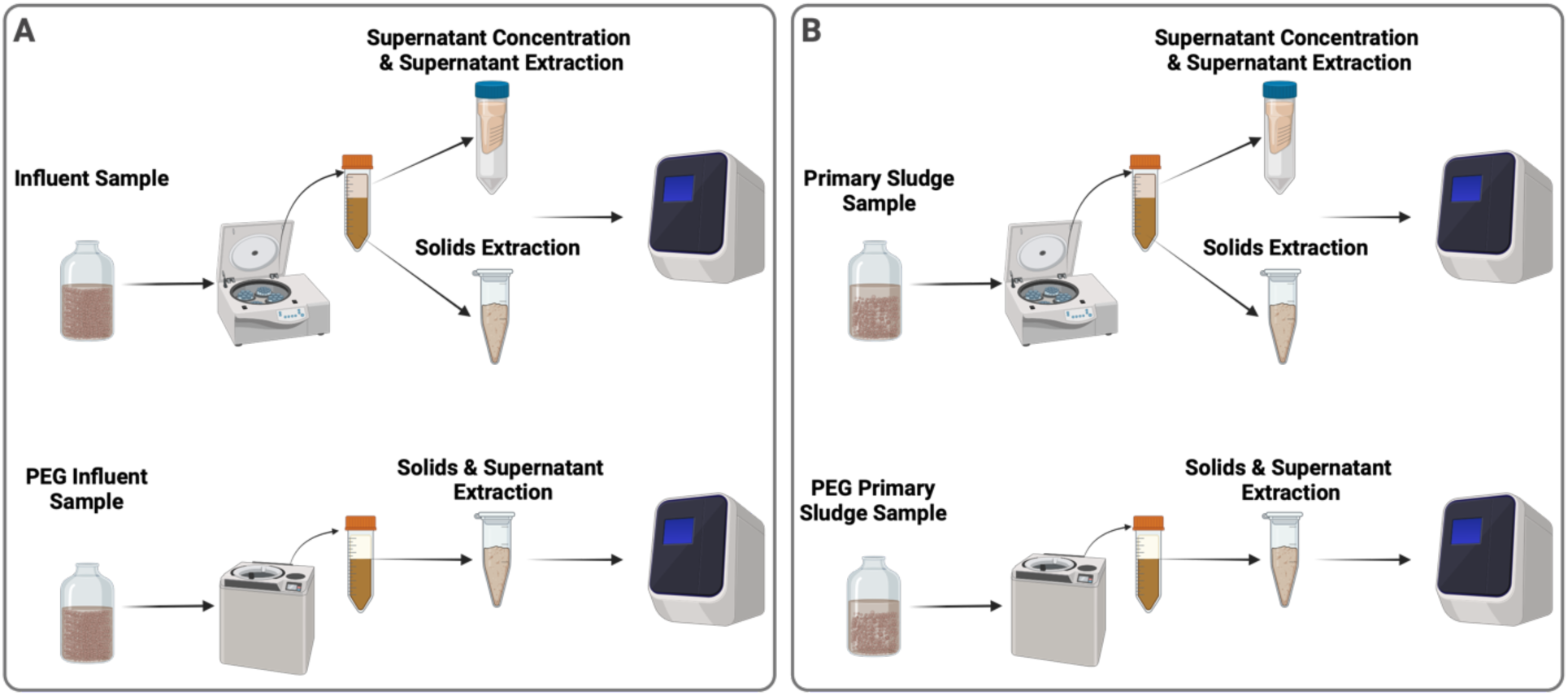
Processing flowcharts of (A) influent samples and (B) primary sludge to identify the partitioning of MTBC and MTB signals within the supernatant and solids with and without PEG addition.

**Table 1.**
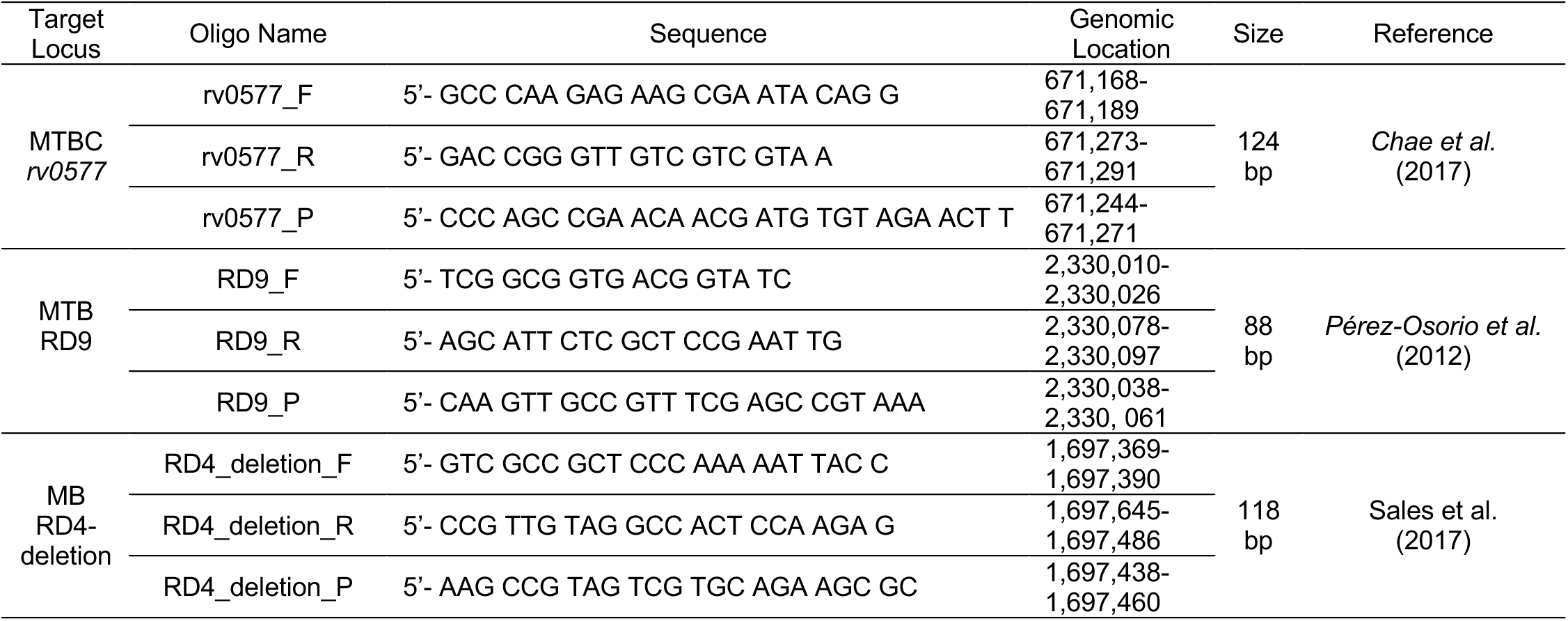
qPCR MTBC, MTB, and MB probe-based assays for TB WEM.

### qPCR Assay Validation and Optimization - Laboratory Propagated Cell Lines

#### DNA Controls

DNA controls were obtained from a panel of laboratory-propagated MTBC cell lines: MTB H37Rv, MB BCG, and a panel of NTM cell lines: *M. kansasii,* and *M. smegmatis.* All cell lines were cultured as previously described in *Madden et al.* (2023) (Madden *et al*., 2023). DNA was extracted following a previously described protocol by *Wong et al.* (2023) (Wong *et al*., 2023).

#### qPCR conditions optimization

The optimal qPCR conditions of the MTBC *rv0577*, MTB RD9, and MB RD4-deletion assays were measured on the CFX96 Real-Time PCR Detection system (BioRad, Hercules, USA) using TaqMan Fast Advanced Master Mix (Applied Biosystems, Foster City, CA) and digital PCR (dPCR) quantified DNA from cultured MTB cells and MB BCG respectively (Table 2). Optimal denaturation durations were determined by running technical triplicates and non-template controls with qPCR reactions conducted at 95°C at 30s, 44 cycles of 95°C for 5, 10, 15, 20, and 30s, and 60°C for 30s. Optimal melting temperatures were determined by running technical triplicates with non-template controls conducted at 95°C at 30s, 44 cycles of 95°C 15s, and annealing temperatures of 0.5°C increments from 57°C to 63°C for 30s. The determined optimal qPCR conditions are comparable to other employed PCR assays for the detection of MTBC, MTB, and MB in clinical and wastewater applications (Wang *et al*., 2019; Lyu *et al*., 2020; Mtetwa *et al*., 2023).

**Table 2.** Optimal qPCR conditions.

| MTBC *rv0577* | MTB RD9 | MB RD4-deletion |
| --- | --- | --- |
| 95°C for 30s | 95°C for 30s | 95°C for 30s |
| 44 cycles: | 44 cycles: | 44 cycles: |
| 95°C for 15s | 95°C for 15s | 95°C for 15s |
| 59°C for 30s | 60°C for 30s | 60°C for 30s |

#### Assay sensitivity

The efficiencies of the MTBC *rv0577*, MTB RD9, and MB RD4-deletion assays were measured using technical triplicates of serially diluted, previously purified, DNA from cultured MTB cells with non-template controls. The DNA was quantified using AbsoluteQ (ABI) dPCR using the AbsoluteQ Universal DNA Digital PCR Master Mix (ABI), MTBC *rv0577*, MTB RD9, and MB RD4-deletion assays, respectively. The reaction efficiencies for MTBC *rv0577*, MTB RD9, and MB RD4-deletion assays are 99.85%, 99.71%, and 99.65%, respectively (data not shown) (Ruijter *et al*., 2013).

#### Assay specificity

The MTBC *rv0577*, MTB RD9, and MB RD4-deletion assays were evaluated for specificity using DNA from the above-mentioned cultured *Mycobacterium spp.* strains. The DNA was quantified using the NanoDrop^TM^ One Microvolume UV Spectrophotometer (ThermoFischer Scientific). MTB was used as a positive control to evaluate the MTBC *rv0577*, MTB RD9 assays, and as a negative control for the RD4-deletion assay. MB BCG was used as a positive control for the MTBC *rv0577* and RD4-deletion assays and a negative control for the MTB RD9 assay. Both *M. kansasii* and *M. smegmatis* were used as negative controls for all three assays, as they belong to the same genus (*Mycobacterium*) but are classified as NTM, because these species do not belong to the MTBC. For each assay, both positive and negative controls were tested at two concentrations, 10 ng/µL (high) and 1 ng/µL (low), using the qPCR conditions as mentioned above. Each test was run in technical triplicates alongside non-template controls to rule out contamination. Positivity was considered when at least two of the three technical replicates showed amplification within 40 qPCR cycles.

### qPCR Assay Validation - Endogenous Targets in Wastewater

#### Locations of collected wastewater with endogenous targets

A panel of MTBC-positive wastewater samples was utilized to validate the herein proposed three new probe-based qPCR assays. The samples include 88 wastewater samples collected from Ottawa, Ontario, Canada, a moderate-risk high-income country metropolitan area (n=70), Mumbai, India, a high-risk middle-income country metropolitan area (n = 2) and a high-risk remote Northern Indigenous community in the Nunangat region (n=16). All locations have reported active TB cases during the respective sampling periods. Ottawa is a moderate-risk, high-income country metropolitan area located in the National Capital Region, where approximately 1 million people reside. The average annual household income in Ottawa is approximately $64,500 Canadian Dollars (CAD) before taxes (Statistics Canada, 2023). Mumbai is a high-risk, middle-income country metropolitan area where approximately 12.5 million people reside and India’s largest city. The average annual household income in Mumbai is approximately ₹230,000 Indian Rupees ($3,778 CAD before taxes)(Rathore, 2024). The Nunangat, Northern Indigenous community is a high-risk remote village located in the Nunavik region of the province of Quebec. Approximately 13 thousand people reside in the Nunavik region across the 14 isolated communities. The average annual household income in the Nunavik region is approximately $35,000 CAD before taxes (Nunavik Statistics Program, 2021).

#### Wastewater collection, transport, and storage prior to analysis

500 mL 24-hour composite primary sludge samples from the city of Ottawa were collected at the Robert O. Pickard Environmental Centre WRRF. This WRRF processes, on average, 436 million litres of wastewater per day, servicing an estimated 1 million residents. Upon collection, the primary sludge samples were immediately refrigerated at 4°C at the WRRF and transported on ice to the University of Ottawa and processed upon receipt. 2.0 L grab samples of influent wastewater and 24-hour composite primary sludge samples from the city of Mumbai were collected at both the Powai and Ghatkhopar WRRFs. These WRRFs process on average, 5.8 and 180 million litres of wastewater per day, servicing an estimated 29 thousand and 900 thousand people, respectively. Upon collection, both the influent wastewater and primary sludge samples were immediately refrigerated at 4°C. The samples were then shipped on ice to the University of Ottawa by express international courier and processed upon receipt. Samples from the Nunangat, Northern Indigenous community were collected from the village’s decentralized wastewater treatment lagoon, where sewage trucks collect wastewater samples from homes and buildings in the village and transport the wastewater to the treatment lagoon daily. Wastewater was collected from the transport trucks using 3D-printed COVID-19 Sewer Cage (COSCa) passive samplers filled with sterile gauze (Hayes *et al*., 2021). COSCa balls were printed using UltiMaker 2+ (Ultimaker) and polylactic acid filament (PLA). Upon collection, the samples were stored at 4°C. The samples were then shipped on ice to the University of Ottawa by express courier and analyzed upon receipt.

#### Wastewater sample nucleic acid extraction

40 mL of homogenized primary sludge samples from Ottawa were centrifuged at 10,000 x g for 45 minutes at 4°C. With the supernatant removed, the pellets were centrifuged again at 10,000 x g for 10 minutes at 4°C. The pellet was stored at -20°C for long-term storage. For retrospective analysis, pellets were thawed overnight at 4°C. Both DNA and RNA were extracted with the AllPrep PowerViral DNA/RNA Kit (Qiagen) using the previously described protocol in Wong *et al*. (2023), D’Aoust *et al*. (2021a), and D’Aoust *et al*. (2021b) and analyzed within 12 hours of DNA extraction (D’Aoust, Graber, *et al*., 2021; D’Aoust, Towhid, *et al*., 2021; Wong *et al*., 2023). 30 mL of influent wastewater settled solids from both Powaii and Ghatkhopar WRRFs were aliquoted. The aliquots were treated with a polyethylene glycol (PEG) 8,000 solution to reach a final working concentration of 80 g/L PEG, 0.3M NaCl using the previously described protocol in Wong *et al*. (2023)(Wong *et al*., 2023). After ultracentrifugation, the resulting pellet was used for both DNA and RNA extraction with the AllPrep PowerViral DNA/RNA Kit (Qiagen) using the previously described protocol in *Wong et al.* (2023), D’Aoust *et al*. (2021a), and D’Aoust *et al*. (2021b) and analyzed within 12 hours of DNA extraction (D’Aoust, Graber, *et al*., 2021; D’Aoust, Towhid, *et al*., 2021; Wong *et al*., 2023). Upon reception of the COSCa samplers used in the Nunangat, Northern Indigenous community, the gauze was removed from the COSCa sampler and mixed with 300 mL of deionized water to elute the collected solids in a clean beaker. The beaker was left to allow the solids to settle for 15 minutes, and 40 mL of the settled solids was pipetted and followed the above primary sludge nucleic acid extraction protocol. All samples were co-extracted with extraction blanks simultaneously using the protocol to rule out contamination in the extraction kits.

#### Wastewater qPCR conditions

All qPCR reactions were run in technical triplicates with non-template controls to ensure accuracy and reliability. A five-point standard curve was prepared using serial dilutions of dPCR-quantified DNA from cultured MTB and MB BCG at concentrations of 100, 40, 20, 5, and 2.5 copies/µL, diluted in molecular-grade water. MTB DNA was used as the control for the MTBC *rv0577* and MTB RD9 assays, while MB BCG DNA was used as the control for the RD4-deletion assay. Inhibition was tested by running pepper mild mottle virus (PMMoV) dilutions at 1/10 and 1/40 with molecular grade RNase-free water. RT-qPCR efficiency ranged from 90-100%, and the coefficient of determination (R^2^) values were greater than 95%. PMMoV is a common fecal matter indicator used in environmental samples (Rosario *et al*., 2009; Haramoto *et al*., 2013). Singleplex, probe-based, single-step RT-qPCR analysis of the PMMoV’s RAP (replication-associated protein) encoding gene was performed, with primers and probes, PCR cycling conditions, and reagent concentrations used for the PMMoV analysis as stated in *Haramoto et al.* (2013), *D’Aoust et al.* (2021a) and *D’Aoust et al.* (2021b) (Haramoto *et al*., 2013; D’Aoust, Graber, *et al*., 2021; D’Aoust, Towhid, *et al*., 2021).

#### Wastewater sample measurements

Measurements of the MTBC *rv0577*, MTB RD9, and MB RD4-deletion targets in wastewater using the herein presented assays are expressed as copies of target DNA per qPCR reaction volume (copies), copies of target DNA per copy per gram of wet wastewater solids (cp/g) and also copies of target DNA per copy per copies of the PMMoV fecal matter indicator (cp/cp).

### Partitioning of Endogenous TB in Municipal Influent Wastewater and Primary Sludge

The partitioning experiments of this study were performed on grab influent wastewater samples and 24-hour composite primary sludge samples from two WRRFs in Mumbai, India. The samples were collected and then shipped on ice via international courier to the University of Ottawa. All samples were subject to the same storage, transport, and holding times prior to the partitioning experiments. The DNA of each fractioned portion of the samples was extracted within 6 hours of performing the partitioning procedure.

To determine the partitioning of endogenous MTBC and MTB in influent wastewater, a 2.0 L grab sample was collected from both Powai and Ghatkopar WRRFs, and each was split into four fractions (n=21): liquid, centrifuged solids, and PEG-precipitated ultracentrifuged solids. MB partitioning experiments were not performed in this study, as the intent of this research is to develop sensitive assays for the measurement of human TB targets in wastewaters. To study influent wastewater partitioning, the samples were processed using the protocol as previously described by *Wong et al.* (2023) and *Mercier et al.* (2022) to produce a total of 5 biological replicates of the liquid fractions, 6 biological replicates of the 0.45-μm filtered solids fraction, 5 biological replicates of the centrifuged solids, 5 biological replicates of the PEG-precipitated ultracentrifuged solids of the samples (Mercier *et al*., 2022; Wong *et al*., 2023). Nucleic acids from the liquid fractions were extracted using the QIAmp Viral RNA Mini Kit (Qiagen) on a QIAcube Connect (Qiagen) automated extraction platform as per the manufacturer’s instructions. Nucleic acids were extracted from the 0.45-μm filtered solids, centrifuged solids, and the PEG-precipitated ultracentrifuged solids fractions using the protocol previously described by *Wong et al. (2023)* and *D’Aoust et al.* (2021) and the AllPrep PowerViral DNA/RNA Kit (Qiagen) (D’Aoust, Towhid, *et al*., 2021; Wong *et al*., 2023). All sample fractions in this portion of the partitioning study were analyzed using probe-based qPCR using the above-mentioned respective reagents and qPCR cycling conditions within 24 hours of nucleic acid extraction.

To determine the partitioning of endogenous MTBC and MTB in primary sludge, a 160mL primary sludge sample was collected from both the Powai and Ghatkopar WRRFs, and each was split into three fractions (n=15): liquid, centrifuged solids, and PEG-precipitated ultracentrifuged solids. To study primary sludge partitioning, the samples were processed using the protocol as previously described by *Wong et al.* (2023) and *Mercier et al.* (2022) to produce a total of 5 biological replicates of the settled solids, 5 biological replicates of the PEG-precipitated ultracentrifuged solids, and 5 biological replicates of the supernatant fractions of the samples (Mercier *et al*., 2022; Wong *et al*., 2023). Nucleic acids from the liquid fractions were extracted using the QIAmp Viral RNA Mini Kit (Qiagen) on a QIAcube Connect (Qiagen) automated extraction platform as per the manufacturer’s instructions. Nucleic acids were extracted from the centrifuged solids and the PEG-precipitated ultracentrifuged solids fractions using the protocol previously described by *Wong et al.* (2023) and *D’Aoust et al.* (2021) and the AllPrep PowerViral DNA/RNA Kit (Qiagen) (D’Aoust, Towhid, *et al*., 2021; Wong *et al*., 2023). All sample fractions in this portion of the partitioning study were analyzed using probe-based qPCR using the above-mentioned respective reagents and qPCR cycling conditions within 24 hours of nucleic acid extraction.

## RESULTS

### Laboratory Propagated Cell Line Validation of qPCR Assays

#### Assay sensitivity

The assay limits of detection (ALOD) and assay limits of quantification (ALOQ) for the MTBC *rv0577*, MTB RD9, and MB RD4-deletion assays were determined using dPCR quantified MTB and MB BCG DNA, respectively (Forootan *et al*., 2017). The purified DNA was serially diluted to concentrations ranging from 1 to 100 copy/μL. The ALOD was determined for MTBC *rv0577* assay, MTB RD9, and MB RD4-deletion at 2.5, 2.5, and 3.2 copies per reaction, respectively, with 95% confidence using conventional methods. The ALOQ was determined for MTBC *rv0577*, MTB RD9, and MB RD4-deletion to be 6.5, 4.5, and 5.4 copies per reaction, respectively. The sensitivity of the probe-based qPCR assays of the herein study using laboratory propagated cell lines are shown to be very similar to SARS-CoV-2, influenza A, influenza B and respiratory syncytial virus sensitivity and hence sufficient for wastewater monitoring (Vogels *et al*., 2020; Mercier *et al*., 2022, 2023).

#### Assay specificity

The MTBC *rv0577* assay detected MTB and MB BCG DNA extracts, while the NTM (*M. kansasii* and *M. smegmatis*) species tested were negative in all the performed tests. The MTB RD9 assay detected DNA extracted from the MTB DNA extract, and all other Mycobacteria DNA extracts tested (MB BCG, *M. kansasii*, and *M. smegmatis*) were negative. The MB RD4-deletion assay only detected the MB BCG DNA extract, and all other Mycobacteria DNA extracts tested (MTB, *M. kansasii* and *M. smegmatis*) were negative (Table 3). As such, the specificity of the probe-based assays of the herein study were validated using laboratory propagated cell lines.

**Table 3.** Validation of assays using laboratory propagated cell lines.

| Organism | qPCR Assay |  |  |
| --- | --- | --- | --- |
|  | MTBC *rv0577* | MTB RD9 | MB RD4-deletion |
| *M. tuberculosis* | + | + | - |
| *M. bovis* BCG | + | - | + |
| *M. kansasii* | - | - | - |
| *M. smegmatis* | - | - | - |

### Endogenous Targets in Wastewaters Validation of qPCR Assays

A panel of 88 wastewater samples was collected from locations with known TB cases: Ottawa, which is a moderate-risk, high-income country metropolitan area (n=70); Mumbai, which is a high-risk middle-income country metropolitan area (n = 2) and a high-risk Nunangat, Northern Indigenous community (n=16) during periods of known ongoing TB cases or outbreaks was used to validate the MTBC *rv0577*, MTB RD9, and MB RD4-deletion assays using endogenous TB target samples. Of the 70 samples from Ottawa, Ontario, Canada, 8 tested positive for both MTBC and MTB (11.4%), and none for MB (0.0%) using the herein proposed MTBC *rv0577*, MTB RD9, and MB RD4-deletion assays (Table 3). All the samples (100%) from Mumbai detected MTBC, MTB, and MB using the proposed MTBC *rv0577*, MTB RD9, and MB RD4-deletion assays. 2 of the 16 samples from the high-risk Nunangat, Northern Indigenous community samples tested positive for MTBC (12.5%), the same 2 were positive for MTB (12.5%), and no samples tested positive for MB using the proposed MTBC *rv0577*, MTB RD9, and MB RD4-deletion assays (Table 4).

**Table 4.** Validation of assays on wastewater samples.

| Sample Location | Sample Date<br>DD/MM/YY | Sample Type | qPCR assays <sup>a</sup> |  |  |  |  |  |  |  |  | Notes on active cases in<br>sampling locations |
| --- | --- | --- | --- | --- | --- | --- | --- | --- | --- | --- | --- | --- |
|  |  |  | MTBC *rv0577* |  |  | MTB RD9 |  |  | MB RD4-deletion |  |  |  |
|  |  |  | copies | cp/g | cp/cp <sup>b</sup> | copies | cp/g | cp/cp | copies | cp/g | cp/cp |  |
| Ottawa | 10/11/21 | WRRF | 0.54 | 70.90 | 4.75x10<sup>-8</sup> | 0.17 | 21.87 | 1.93x10<sup>-6</sup> | nd | nd | nd | 61 confirmed cases in Ottawa in 2021 |
| Ottawa | 27/11/21 | WRRF | 0.93 | 120.17 | 8.12x10<sup>-8</sup> | 1.07 | 138.34 | 1.21x10<sup>-5</sup> | nd | nd | nd |  |
| Ottawa | 04/12/21 | WRRF | 0.49 | 63.89 | 4.16x10<sup>-8</sup> | 0.07 | 8.65 | 3.75x10<sup>-7</sup> | nd | nd | nd |  |
| Ottawa | 19/04/22 | WRRF | 0.86 | 110.25 | 1.39x10<sup>-7</sup> | 0.13 | 16.45 | 9.38x10<sup>-7</sup> | nd | nd | nd | 59 confirmed cases in Ottawa in 2022 |
| Ottawa | 13/06/22 | WRRF | 0.12 | 15.86 | 1.53x10<sup>-7</sup> | 0.11 | 14.28 | 6.39x10<sup>-6</sup> | nd | nd | nd |  |
| Ottawa | 17/06/22 | WRRF | 0.69 | 89.60 | 1.19x10<sup>-7</sup> | 0.19 | 24.91 | 1.14x10<sup>-6</sup> | nd | nd | nd |  |
| Ottawa | 04/07/22 | WRRF | 0.38 | 49.16 | 9.45x10<sup>-8</sup> | 0.18 | 23.23 | 2.04x10<sup>-6</sup> | nd | nd | nd |  |
| Ottawa | 15/11/22 | WRRF | 0.36 | 47.14 | 1.58x10<sup>-8</sup> | 0.15 | 19.59 | 8.05x10<sup>-7</sup> | nd | nd | nd |  |
| Mumbai - P | 12/07/23 | WRRF | 12622.98 | 1.68x10<sup>6</sup> | 4.34x10<sup>0</sup> | 5012.99 | 6.68x10<sup>5</sup> | 1.73x10<sup>0</sup> | 2.46x10<sup>2</sup> | 3.27x10<sup>4</sup> | 8.47x10<sup>-2</sup> | An estimate of 67 000 cases of active TB in Mumbai in 2023 |
| Mumbai - G | 13/07/23 | WRRF | 8313.63 | 1.42x10<sup>6</sup> | 1.22x10<sup>0</sup> | 2843.98 | 3.79x10<sup>5</sup> | 0.39x10<sup>0</sup> | 6.92x10<sup>1</sup> | 9.20x10<sup>3</sup> | 2.42x10<sup>-2</sup> |  |
| Nunangat | 14/02/23 | Sewage Truck | 0.73 | 93.98 | 1.98x10<sup>-4</sup> | 0.16 | 20.87 | 5.64x10<sup>-3</sup> | nd | nd | nd | 81 confirmed cases in the Nunangat, Northern Indigenous community in 2023 |
| Nunangat | 28/02/23 | Sewage Truck | 1.60 | 206.53 | 2.40x10<sup>-3</sup> | 0.34 | 43.39 | 6.5x10<sup>-2</sup> | nd | nd | nd |  |

<sup>a</sup> The cycle when the fluorescence signal crosses the threshold (C<sub>T</sub>) for each positive sample is shown. nd, not detected. Each reaction mixture contained 1X TaqMan™ Fast Advanced Master Mix (with UNG) (Biosystems, Foster City, CA), 500 nmol of each primer, 250 nmol of each TaqMan probe, and 3 µL of template DNA. Thermal cycling conditions for the BioRad CFX96: one cycle of 95°C for 30 s; 44 cycles of 95°C for 15 s, and 59°C for 30 s for MTBC (*rv0577*) assay, one cycle of 95°C for 30 s; 44 cycles of 95°C for 15 s, and 60°C for 30 s for MTB (RD9) and MB (RD4-deletion) assays.
<sup>b</sup> The number of copies normalized for PMMoV
Mumbai - P: Powai WRRF
Mumbai - G: Ghatkhopar WRRF

Cases of active TB in Ottawa fluctuate between 40 to 70 cases reported per year in the city of approximately 1M people, with more than half being cases of pulmonary TB (Ottawa Public Health, no date; Public Health Ontario, 2024). The city’s public health unit, Ottawa Public Health reported 61 cases of active TB in 2021 and 59 cases in 2022, resulting in incidences of 5.8 and 5.5 per 100,000 people, respectively. This is observed in the detection of MTBC and MTB using the proposed assays in primary sludge samples collected from Ottawa (Table 4).

In 2022, an estimated 354 million people in India were presumed to have active TB infection, the highest burden globally (Chauhan *et al*., 2023). In addition, Mumbai experienced approximately 67,000 cases of active TB, resulting in an incidence of 313 cases per 100,000 people in 2022, which is reflected in the detection and high concentrations of MTBC and MTB using the proposed assays (Shah *et al*., 2024). The high detection rate of MB (100%) in Mumbai likely reflects the endemic nature of MB in the region, where close human-animal interactions and agricultural practices may occur (Table 4) (Ramanujam and Palaniyandi, 2023).

In 2023, the Nunangat, Northern Indigenous community’s public health unit, the Nunavik Regional Board of Health and Social Services reported 81 cases of active TB in Nunavik (Madalyn Howitt, 2024). This equates to an estimated incidence of 643 reported cases per 100,000 people in the Nunavik region. These reported cases highlight the disproportionate burden of TB in Indigenous communities compared to that of non-Indigenous, Canadian-born individuals (0.3 per 100 000) (Public Health Agency of Canada, 2024). Despite this higher incidence, the MTBC and MTB measurement concentrations in wastewater samples from the Nunangat, Northern Indigenous community were similar to those observed in Ottawa. This similarity may reflect that the 81 reported cases in Nunavik were distributed across 14 villages, potentially with significant lower case numbers in the specific location where we collected samples. Additionally, the case count at the time of sampling, when there was not a major outbreak TB identified, may have been low at this specific sampling location, leading to lower concentrations detected in the samples. Nonetheless, the detection of the MTBC and MTB signals using the proposed assays confirms the utility of WEM for TB in this Nunangat, Northern Indigenous community, as well as other remote and underserved communities (Table 4).

### Partitioning of Endogenous TB in Municipal Influent Wastewater and Primary Sludge

Endogenous partitioning experiments for MTBC and MTB were conducted on influent wastewater and primary sludge samples collected from two WRRFs in Mumbai, India where TB measurement magnitudes were sufficient to fractionate the samples. These experiments employed the proposed probe-based MTBC *rv0577* and MTB RD9 assays. MB RD4-deletion was not tested in the partitioning portion of this study as the herein study is focussed on the development of wastewater monitoring assays for human TB. In influent wastewater, 86.4 ± 15.7% of the total MTBC and 97.3 ± 2.4% of the total MTB signal was detected in the PEG-precipitated ultracentrifuged solids fraction (Bonferroni adjusted p = 0.49) (Jafari and Ansari-Pour, 2018). The centrifuged solids fraction accounted for 31.0 ± 1.6% of the MTBC signal and 14.8 ± 1.4% of the MTB signal (adj. p = 0.00006). In contrast, the liquid fraction contained <0.1% of the total signal for MTBC, and no signal was detected for MTB (adj. p = 0.045) (Figure 4). No measurement of both MTBC and MTB was observed in the 0.45-μm filtered solids of the influent wastewater. Note that the above partitioned percentages of MTBC compared to MTB in influent wastewater are not statistically distinct from one another, except for the centrifuged solids fraction. This distinction in centrifuged solids fraction particitioning may be attributed to the heterogeneity in sample composition, as the dispersed nature of influent wastewater solids could influence their partitioning behaviour. Overall, these findings indicate that the vast majority of MTBC and MTB in influent wastewater partitions to the wastewater solids and is further enhanced with the addition of PEG-precipitation compared to the liquid extraction method.

**Figure 4.**
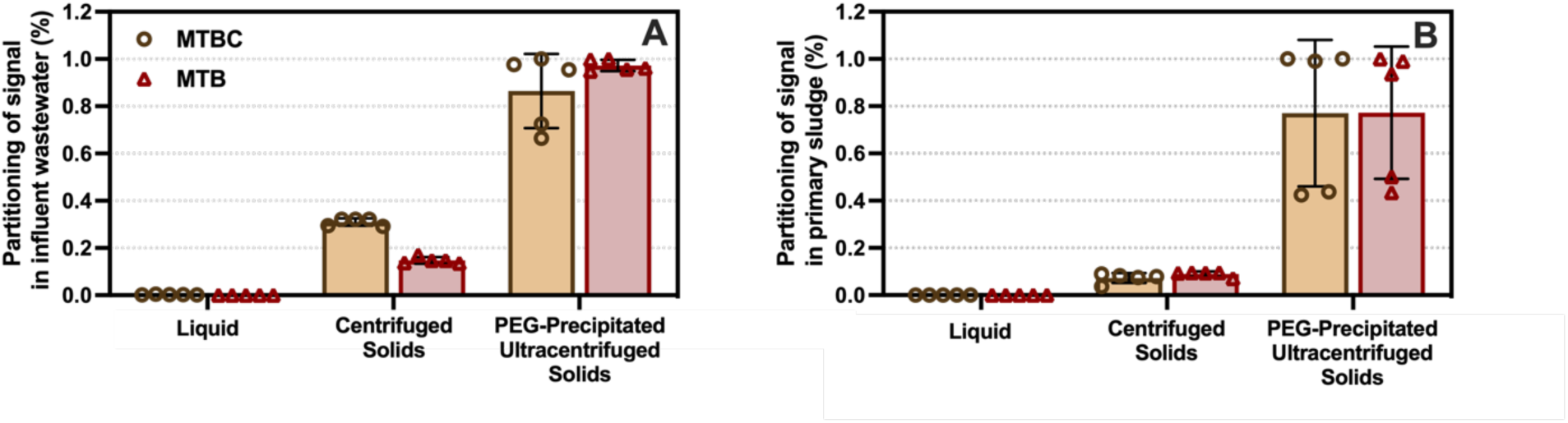
Partitioning of measured endogenous MTBC and MTB signal present in (A) influent wastewater (n=2) and (B) primary sludge (n=2). Mean percentages and standard deviations are displayed. Where the standard deviation is too small, the error bars are not displayed.

In primary sludge, most of the MTBC and MTB signal was found in the PEG-precipitated ultracentrifuged solids fraction. Specifically, 77.0 ± 31.0% of the total MTBC and 77.3 ± 28.0% of the total MTB signals were detected in this fraction (adj. p = 0.99). The centrifuged solids accounted for 7.2 ± 2.1% of the MTBC signal and 9.0 ± 1.1% of the MTB signal (adj. p = 0.07). In contrast, the liquid fraction contained <0.1% of the total signals for both MTBC and MTB (adj. p = 0.43) (Figure 4). Again, it is noted that all above partitioned percentages of MTBC compared to MTB in primary sludge are not statistically distinct from one another. These findings again indicate that the majority of MTBC and MTB in primary sludge partitions to the solids and is further enhanced with the addition of PEG-precipitation compared to the liquid extraction method. These results highlight the importance of targeting the solids fraction of wastewaters during sample collection and again during sample processing to enrich and optimize the recovery of MTBC and MTB markers. While the solids-based extraction method is effective, the addition of PEG-precipitation using ultracentrifugation is particularly effective for concentrating MTBC and MTB signals in both influent wastewater and primary sludge, which can be used to guide sample enrichment and augment detection sensitivity in WEM applications, especially during periods of low signal.

### Contextualization of TB WEM in Nunangat, Northern Indigenous Community Wastewaters as an Early Warning System and to Monitor Transmission and Recovery

The Nunangat, Northern Indigenous community described in this study experiences a disproportionate burden of TB compared to the non-Indigenous, Canadian-born population. Indigenous communities in Canada, and more specifically, Inuit populations, consistently face TB incidences that are several hundred times than the national average, driven by a combination of socio-economic, institutional, and historical factors. Overcrowded housing, food insecurity, limited access to healthcare, geographic isolation, and systemic inequalities and inequities in social determinants of health contribute significantly to this disparity (Kilabuk *et al*., 2019). This burden is exasperated by a complex medical history, which includes forced treatment and relocation by the Government of Canada originating from forms of colonialism (Allan and Smylie, 2015). As a result, many Indigenous and Inuit individuals are hesitant to obtain medical diagnosis or treatment for TB (Jetty, 2020). In this context, TB WEM offers an alternative for TB monitoring in these communities where there are historical and ongoing barriers toward medical diagnoses. Detecting and quantifying TB in wastewater allows for non-invasive and anonymous monitoring, avoiding the need for direct individual participation (Doorn, 2022). This approach respects privacy and minimizes potential stigmatization, which is particularly important in communities where TB is not only a public health issue but also a stigmatized issue. Additionally, WEM provides a population-level snapshot of TB prevalence, offering insights into outbreaks without clinical data (Wong *et al*., 2023). In Indigenous communities, especially remote communities, access to healthcare services is often limited. The herein proposed assays and the partitioning results provide an approach that can hence serve as an early warning system, identifying trends in transmission and trends in recovery. It is necessary, however, to clearly state that any adoption of TB WEM in Northern Indigenous communities must occur in collaboration with the Indigenous communities themselves. This adoption requires culturally sensitive and respectful approaches. Monitoring strategies and techniques must be co-developed with community members and stakeholders to ensure that the methodology and goals align with the community’s values and priorities. By incorporating Indigenous knowledge and perspectives into the research process, WEM can become a valuable tool to reduce TB transmission in priority populations rather than a perpetuation of historical trauma.

## CONCLUSION

This study presents the development and validation of three probe-based qPCR assays for the detection of MTBC, MTB, and MB in wastewater, offering a sensitive and specific tool for TB monitoring. The assays were validated using propagated cell lines and wastewater samples from three geographically and socioeconomically diverse locations with confirmed TB infections: Ottawa, Canada; Mumbai, India; and a remote Northern Indigenous community in Nunangat, Canada. These settings represent a spectrum of TB incidence rates and infrastructure contexts, demonstrating the versatility and applicability of the assays in both high- and low-burden areas. The partitioning of MTBC and MTB in influent wastewaters and primary sludge both demonstrated strong partitioning to the solids fraction of wastewaters and recovery is further enhanced with the addition of PEG-precipitation and ultracentrifugation. By offering a non-invasive, anonymous, and cost-effective approach to monitoring TB at a population level, these assays and the partitioning information provides a new monitoring tool that addresses key challenges in traditional monitoring methods. This is particularly valuable for remote, underserved, priority communities, where healthcare infrastructures may be limited and difficult to obtain. Overall, this study highlights the potential of a WEM approach to address global health challenges, emphasizing the critical role of developing innovative solutions for TB control and informed public health interventions.

## Ethical considerations

Before commencing this research, the authors received advice from the research ethics board of the University of Ottawa and the Canadian Research Ethics Board. The clinical data used in this study was gathered and consolidated by local public health units, adhering to applicable regulations and guidelines, and was entirely anonymous.

## Acknowledgements

The authors wish to acknowledge the help and assistance of the community members in Nunavik and the Nunavik Regional Board of Health and Social Services and all their employees involved in the project. The authors wish to acknowledge Katie Madden from the Sun Laboratory, University of Ottawa, for preparing and providing the cell cultures used in this study as positive controls. The authors would like to acknowledge funding from the CIHR Applied Public Health Chair in Environment, Climate Change and One Health, awarded to Dr. Robert Delatolla.

## Author contributions statement

R.D. conceived the experiments, T.B.N. and S.W. conducted the experiments, T.B.N., E.M, and R.D. analyzed the results, T.B.N., E.M., N.H., P.K., S.W., and R.D. edited the manuscript. All authors reviewed the manuscript.

## Competing interest statement

The authors declare that no known competing financial interests or personal relationships could appear to influence the work reported in this manuscript.

## Data availability statement

Data will be available upon reasonable request by contacting the corresponding author.

## Funding

This research was supported by the CIHR Applied Public Health Research Chair in Environment, Climate Change and One Health, awarded to Dr. Robert Delatolla. The funding had no involvement in the study design, data collection, data analysis, data interpretation, nor the writing or decision to submit the paper for publication.

## Notes

### Competing Interest Statement

The authors have declared no competing interest.

### Author Declarations

The study used only openly available human data that were originally located at: https://www.publichealthontario.ca/-/media/Documents/T/24/tuberculosis-ontario-epi-summary-2019-2023.pdf?rev=c6ff541604bd4e6491aa7cbb97944439&sc_lang=en https://doi.org/10.3389/FTUBR.2024.1454277 https://nunatsiaq.com/stories/article/nunavik-health-board-says-tuberculosis-present-in-8-communities/

